# Multicenter analysis of atrioesophageal fistula rates before and after adoption of active esophageal cooling during atrial fibrillation ablation

**DOI:** 10.1101/2023.02.21.23286267

**Authors:** Javier Sanchez, Christopher Woods, Jason Zagrodzky, Jose Nazari, Matthew Singleton, Amir Schricker, Annie Ruppert, Babette Brumback, Benjamin Jenny, Charles Athill, Christopher Joseph, Dipak Shah, Gaurav Upadhyay, Erik Kulstad, John Cogan, Jordan Leyton-Mange, Julie Cooper, Kamala Tamirisa, Samuel Omotoye, Saroj Timilsina, Alejandro Perez-Verdia, Andrew Kaplan, Apoor Patel, Alex Ro, Andrew Corsello, Arun Kolli, Brian Greet, Danya Willms, David Burkland, Demetrio Castillo, Firas Zahwe, Hemal Nayak, James Daniels, John MacGregor, Matthew Sackett, Michael Kutayli, Michel Barakat, Robert Percell, Spyridon Akrivakis, Steven C. Hao, Taylor Liu, Ambrose Panico, Archana Ramireddy, Daniel Benhayon Lanes, Edward Sze, Greg Francisco, Jose Silva, Julia McHugh, Kai Sung, Leon Feldman, Nicholas Serafini, Raymond Kawasaki, Richard Hongo, Richard Kuk, Robert Hayward, Shirley Park, Andrew Vu, Christopher Henry, Shane Bailey, Steven Mickelsen, Taresh Taneja, Westby Fisher, Mark Metzl

## Abstract

**Background:** Active esophageal cooling reduces the incidence of endoscopically identified severe esophageal lesions during radiofrequency (RF) catheter ablation of the left atrium for the treatment of atrial fibrillation. No atrioesophageal fistula (AEF) has been reported to date with active esophageal cooling, and only one pericardio-esophageal fistula has been reported; however, a formal analysis of the AEF rate with active esophageal cooling has not previously been performed.

**Methods:** Atrial fibrillation ablation procedure volumes before and after adoption of active cooling using a dedicated esophageal cooling device (ensoETM, Attune Medical) were determined across 25 hospital systems with the highest total use of esophageal cooling during RF ablation. The number of AEFs occurring in equivalent time frames before and after adoption of cooling were then determined, and AEF rates were compared using generalized estimating equations robust to cluster correlation.

**Results:** Throughout the 25 hospital systems, which included a total of 30 separate hospitals, 14,224 patients received active esophageal cooling during RF ablation, with the earliest adoption beginning in March 2019 and the most recent beginning in March 2022. In the time frames prior to adoption of active cooling, a total of 10,962 patients received primarily luminal esophageal temperature (LET) monitoring during their RF ablations. In this pre-adoption cohort a total of 16 AEFs occurred, for an AEF rate of 0.146%, in line with other published estimates of <0.1% to 0.25%. No AEFs were found in the cohort treated after adoption of active esophageal cooling, yielding an AEF rate of 0% (P<0.0001).

**Conclusion:** Adoption of active esophageal cooling during RF ablation of the left atrium for the treatment of atrial fibrillation was associated with a significant reduction in AEF rate.

## Introduction

The treatment of atrial fibrillation (AF) using pulmonary vein isolation (PVI) involves risks to collateral structures, including the esophagus.[1-4] Esophageal injuries include ulceration, hematoma, spasm, disorders of esophageal motility, and atrioesophageal fistula (AEF). [5-9] AEF has a high mortality rate, with the latest data suggesting a survival rate of 64.7% with surgery, 31.0% with endoscopic repair, and 2.8% with medical therapy alone.[10] Despite the use of temperature sensors for luminal esophageal temperature (LET) monitoring, esophageal deviation devices, or technological advances such as contact force measuring catheters, the incidence of AEF does not appear to be decreasing,[11-13] with incidence rates for AEF estimated to range from 0.1% to 0.25%,[2] but possibly higher.[14] These estimates may reflect underreporting the true incidence as well.[10]

Methods used to mitigate the risk of AEF include reducing ablation energy applied to the left atrium posterior wall, monitoring esophageal temperature [15-17], and mechanically displacing the esophagus.[18] Despite the long-standing uncertainty of its benefits [15-17, 19, 20] and the recent clinical studies demonstrating either no benefit or trends towards harm with its use, LET monitoring is still widely used.[21-23] Active esophageal cooling using a dedicated esophageal cooling device is a newer technique that has shown benefits in preclinical studies, mathematical modeling, and clinical studies.[24-34] An international multicenter randomized controlled trial is currently underway (NCT04577859), but this study relies on the surrogate endpoint of endoscopically detected esophageal lesion (EDEL) reduction. Because an event rate of <1% requires an extremely large sample size, no study to date has evaluated the effectiveness of any strategy in reducing AEFs. We therefore sought to perform the first study of active esophageal cooling sufficiently powered to determine the efficacy of this technique in reducing AEF formation.

## Methods

### Study Design

This was a retrospective review of data from 25 hospital systems to quantify the effect of active esophageal cooling by (1) measuring AEF rates across hospital systems with the highest use of this method, and then (2) comparing these rates before and after the adoption of active esophageal cooling. The 25 hospital systems included a total of 30 separate hospitals. With central IRB review and additional approvals as required by each hospital site, data were extracted by site investigators to determine the total number of RF catheter ablations performed for the treatment of atrial fibrillation over the study time frame and the number of AEFs that occurred over this time frame. Methods used to determine these data included review of electrophysiology lab registries, the Electronic Medical Record, other recorded sources of patient follow-up (cardiology clinic records, hospital-based registry, e.g.), device shipment records, and inquiries to physician operators and laboratory managers.

### Study Time Frame

The time frame of review at each site was based on the date of adoption of active esophageal cooling, such that an equivalent time before and after adoption was analyzed. The cutoff for observation across all sites was December 2022, such that the time frame of observation was from adoption forward to December 2022 and from adoption backward for an equivalent number of months. For example, in the case of a site adopting esophageal cooling into practice in May 2021, the time frame studied included a minimum of 38 months, with the 19 months from May 2021 to December 2022 consisting of the cohort of patients treated with active esophageal cooling, and the equivalent time frame of 19 months prior to adopting active esophageal cooling into practice consisting of the cohort of patients treated with alternative esophageal protective strategies (primarily LET monitoring). Data from each site were obtained by electrophysiologist physicians, research assistants, and cardiology or electrophysiology fellows.

### Study Population

Patients undergoing RF catheter ablation for the treatment of AF over the time frame of analysis across 25 hospital systems were included in this study. Most hospital systems consisted of a single hospital, three systems consisted of two hospitals, and one system consisted of three hospitals at which RF catheter ablations were performed. The 25 hospital systems that had the highest number of cases completed with active esophageal cooling (irrespective of when adoption occurred) were determined from data obtained from the manufacturer of the esophageal cooling device and verified by hospital staff on each site.

### Study Device

In all patients treated with active esophageal cooling, the cooling was achieved with a dedicated esophageal cooling device (ensoETM, Attune Medical, Chicago, IL). This device is a multi-channel silicone tube that is inserted into the esophagus in a manner similar to a standard orogastric tube (Figure 1). The device is connected to a temperature-controlled heat-exchanger that circulates water through the closed-loop system of the device. This makes it possible to control the temperature of the esophagus by adjusting the temperature of the circulating water. For cooling applications, a water temperature of 4 °C is used. The device replaces the standard orogastric (OG) tube typically placed in patients during anesthesia and replaces the temperature probe that would be placed to perform LET monitoring.

**Figure 1.**
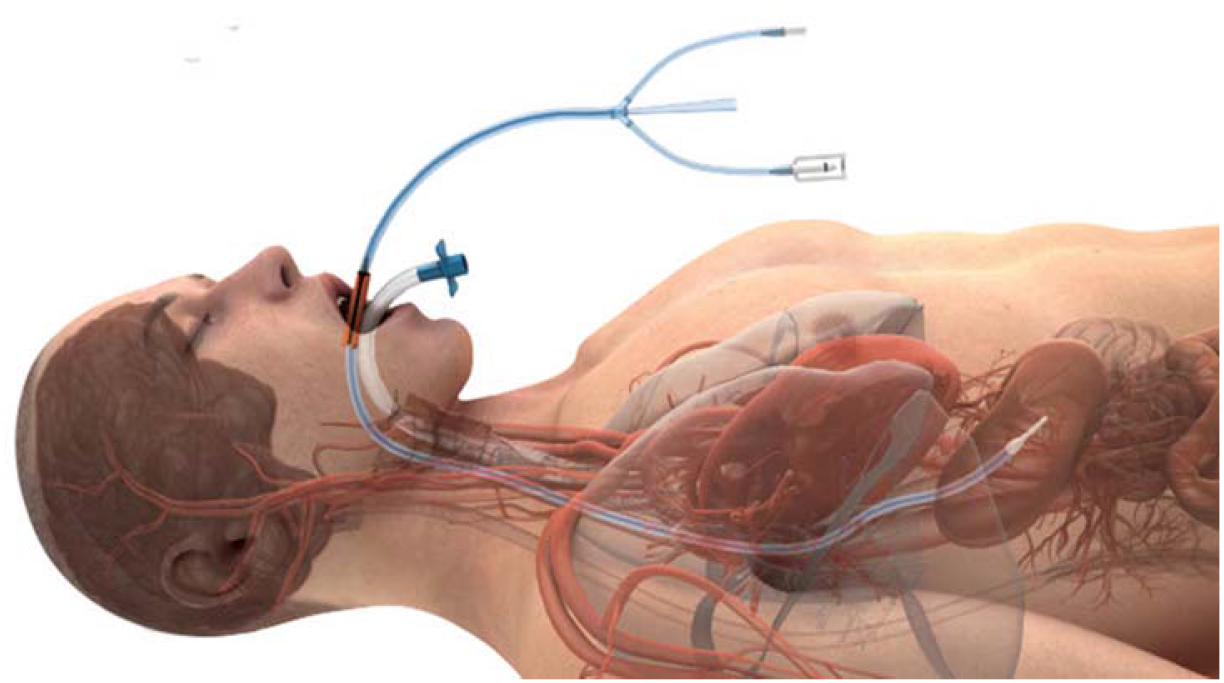
An active esophageal cooling device shown in place in the esophagus, with endotracheal tube shown in place in the trachea (with permission).

### Statistical Analysis

AEF event rates were determined by quantifying at each site the total number of patients treated over the pre- and post-cooling time periods. These event rates were then compared by using a risk difference computed with the use of generalized estimating equations with a linear link and independent working correlation structure. This targets overall rates that weight medical centers by caseload and adjusts for correlation of pre- and post-measures. The null hypothesis is that the risk difference is zero, which was tested with a two-sided level 0.05 hypothesis test. The estimated risk difference compares the AEF event rate in the pooled treatment (post-adoption) group with the AEF event rate in the pooled control (pre-adoption) group; the generalized estimating equations methodology adjusts the standard error of this estimate for possible correlation within medical centers of pre- and post-measures. A sensitivity analysis was then conducted by assuming missed AEF events and determining the impact of missed events on the statistical significance of the difference between groups. For this analysis the most conservative approach was taken, with all missing events assumed to come from the cohort of patients treated with active esophageal cooling.

## Results

### Site Characteristics

The earliest adoption of active esophageal cooling across the 25 hospital systems occurred in September 2018 and the most recent in March 2022. Procedure volumes for RF catheter ablation of the left atrium at each hospital system ranged from 6 to 44 per month, with a mean procedure volume per month of 19.4 ± 10.7 (Figure 2).

**Figure 2.**
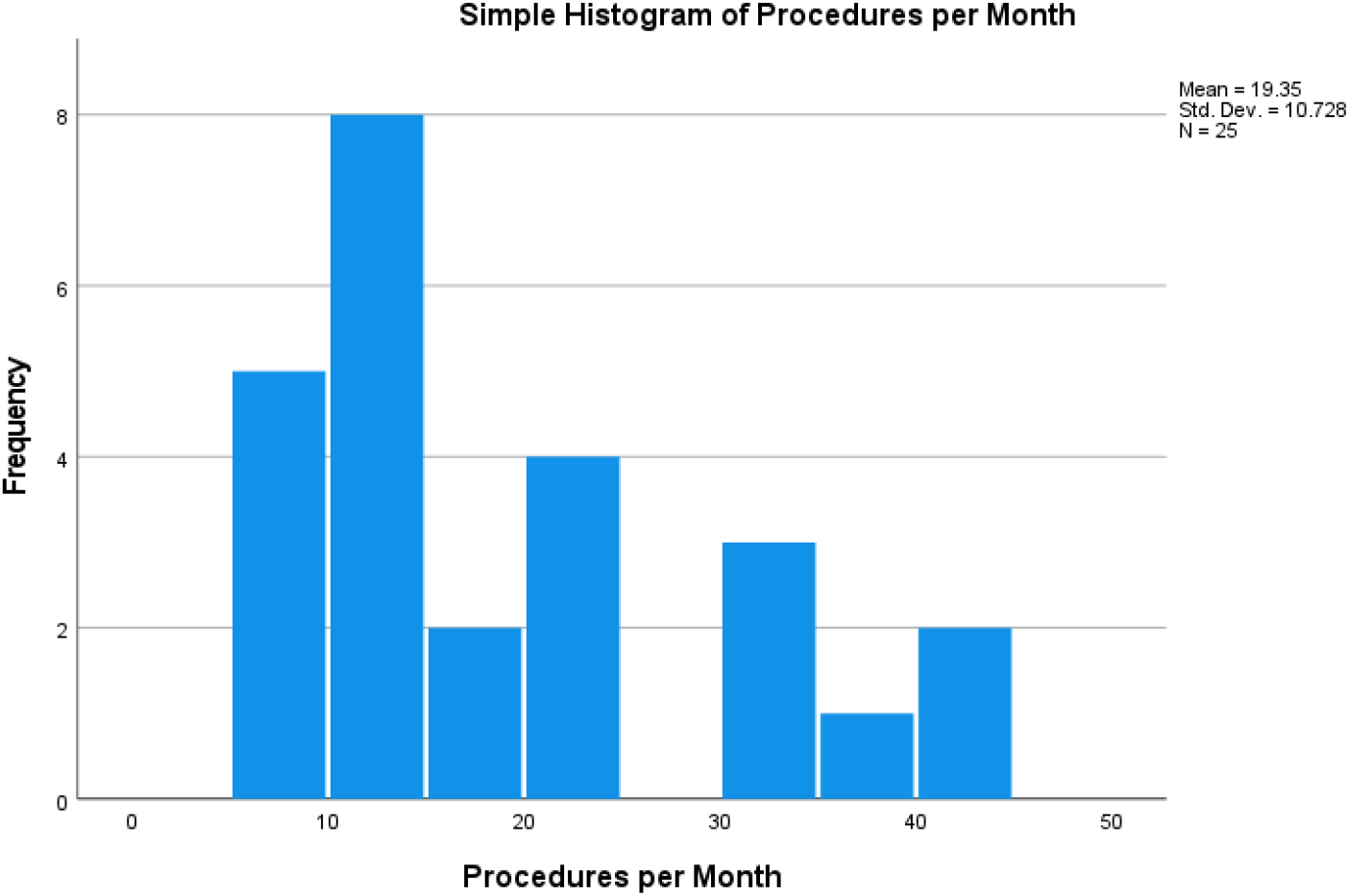
Histogram of procedure volume per month at each hospital system. Bar height represents the number of sites performing the procedure volume per month represented by position on the x-axis.

### Procedure Volumes

The number of patients treated with active esophageal cooling at each hospital system ranged from 212 to 1700. In total, 14,224 patients were treated with active esophageal cooling during RF catheter ablations across the 25 hospital systems over the post-adoption time frame. In the time frame prior to adoption of active cooling, a total of 10,962 patients received RF catheter ablation for the treatment of AF. Most of these patients received LET monitoring during their RF ablations, using either single or multi-sensor temperature probes; a smaller subset also received esophageal deviation.

### Procedure Characteristics

The general procedures utilized by operators at the study sites reflect those of most centers performing left atrial catheter ablation in the United States. Patients were treated under general anesthesia for their ablation procedure. Electrophysiologists performed primarily wide area circumferential pulmonary vein isolation with additional posterior wall isolation, as well as mitral isthmus lines and cavotricuspid isthmus lines dependent on physician practice. Anticoagulation was administered with a heparinized target activated clotting time typically ranging from 300 to 350 seconds. Most sites utilized either the CARTO^®^ mapping system (Biosense Webster, Inc., Diamond Bar, CA) or the EnSite Precision™ cardiac mapping system (Abbott, Abbott Park, IL) to obtain electroanatomical maps and create a three-dimensional geometry. Intracardiac echocardiography was used by almost all sites, as was an irrigated ablation catheter with contact force sensing, such as the ThermoCool^®^ SmartTouch^®^ Surround Flow (STSF) catheter (Biosense Webster, Inc., Diamond Bar, CA) or the TactiCath™ Contact Force Ablation catheter (Abbott, Abbott Park, IL). Power settings ranged from 30 W to 50 W, with most operators (approximately 70%) utilizing high power short duration settings (40 W to 50 W) for ablation. When utilized, a Visitag Surpoint^®^ ablation index (Biosense Webster, Inc.) of 350 to 400 units on the posterior wall, and 450 to 550 units on the anterior wall, lateral wall, and septum was targeted. Similarly, a Lesion Index (LSI) between 4.5 and 5 posteriorly, and between 5.5 and 6 anteriorly was generally targeted. No specific changes of equipment or ablation technique at the time of adoption of esophageal cooling were reported by sites; however, a number of operators report performing more frequent posterior wall ablation after adopting cooling.

### AEF Events

In the cohort of patients treated across the 25 systems prior to the adoption of active esophageal cooling, a total of 16 AEFs occurred, yielding an AEF rate of 0.146%. In the cohort of patients treated after adoption of active esophageal cooling, no AEFs were identified, representing an AEF rate of 0%, and a statistically significant difference between groups (P<0.0001). Figure 3 depicts these results, with the middle column showing the date of adoption for each site, each row representing one of the 25 different hospital systems, and the length of the horizontal bars representing the number of months before and after adoption of active esophageal cooling. At the left and right ends of each bar is a numerator of the number of AEFs and denominator of the number of RF catheter ablations performed. Each “x” represents the approximate time of occurrence of an AEF.

**Figure 3.**
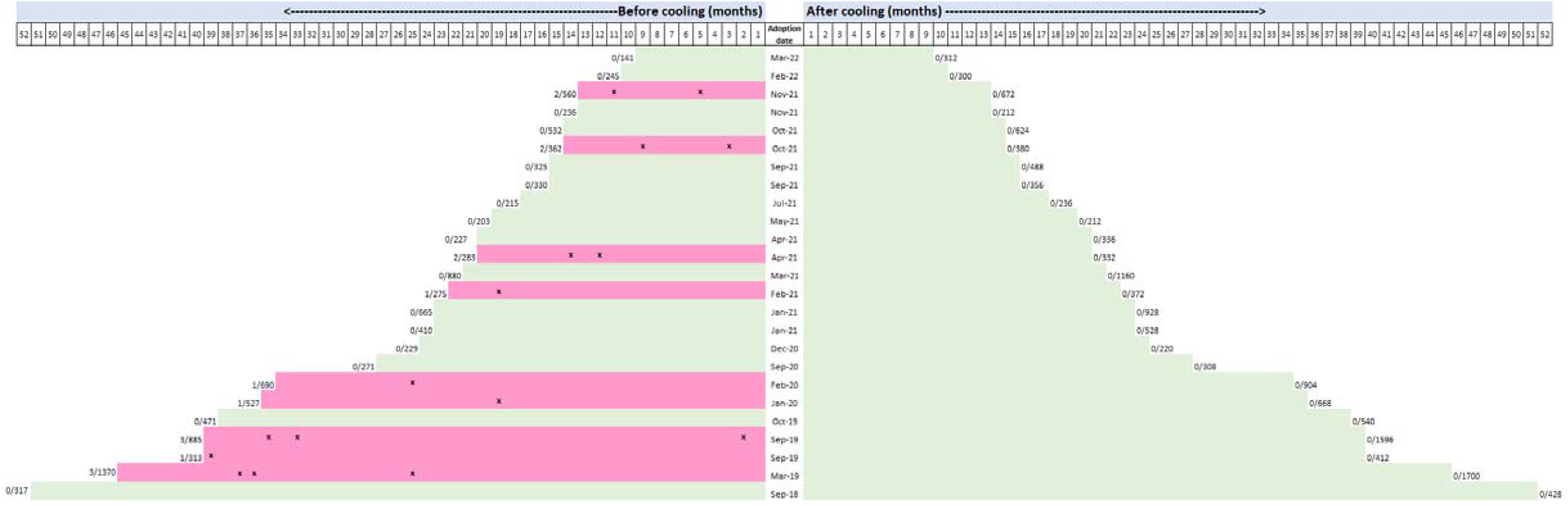
Graphical representation of AEF rates. Middle column shows the date of adoption for each site, with each row representing one of the 25 different hospital systems, and the length of the horizontal bars representing the number of months before and after adoption of active esophageal cooling. At the left and right ends of each bar is the numerator of the number of AEFs and the denominator of the number of RF catheter ablations performed, with “x” marks indicating the approximate time each AEF occurred. Abbreviation: AEF - atrioesophageal fistula.

### Sensitivity Analysis

A sensitivity analysis was conducted, adding missed AEFs only to the cohort of patients treated with active esophageal cooling (the most conservative approach). Using our generalized estimating equations approach with an independent working correlation structure, the point estimate of the risk difference remains the same no matter to which hospital systems we add the missed AEFs. However, the standard error for the comparison would be larger if the added AEFs were concentrated in a single site, or in a smaller number of sites, than if they were spread across all the sites. Furthermore, it would be larger if the added AEFs were in sites with smaller caseloads. Statistical significance using a threshold for type I error of 0.05 is maintained upon adding up to 5 missed AEFs at a single site (large or small) or up to 9 missed AEFs spread across the largest sites.

## Discussion

In this largest study to date on the effectiveness of any technique to reduce the incidence of AEF after left atrial ablation for the treatment of AF, we found that the adoption of active esophageal cooling was associated with a large clinically and statistically significant decrease in AEF rates. This is also the first study to systematically estimate AEF rates using more rigorous methods than have previously been employed, overcoming the limitations inherent in anonymous surveys and avoiding the constraints of existing large databases. Large databases such as the National Inpatient Sample (NIS), the Nationwide Readmissions Database (NRD), and the National Hospital Ambulatory Medical Care Survey (NHAMCS) are limited by reliability concerns and the lack of specific diagnosis codes for AEF.[5, 35] In contrast, our study relied on actual operators and hospital staff reviewing their own hospital system data, and involved a more detailed analysis than previously attempted, utilizing a population of over 25,000 ablation cases.

Preclinical data as well as mathematical modeling support the findings of a significant effect size, with a dose-response effect of coolant temperature shown in a large animal model,[24] and a significant reduction of lethal isotherm formation in the esophagus shown with mathematical models.[33, 36] Although randomized clinical studies have shown reductions in severe esophageal lesions with active esophageal cooling,[31, 37, 38] the effect sizes seen were not such that the findings of no AEF in our large population sample would be expected. Downstream effects on inflammatory markers (which are well-described in the burn literature) [39-41] may be another mechanism involved in reducing the likelihood of fistula formation with cooling. Insulating effects from the pericardial fat, fibrous pericardium, and serous layers minimize cooling in the atrial myocardium so that effective lesions can still be placed,[33] with long-term follow up data confirming no decrease in freedom from atrial arrhythmias at one year with active esophageal cooling compared with LET monitoring.[32] A larger volume of retrospective data further suggests improvement in freedom from arrhythmias with cooling,[42] which may be due to differences in lesion placement sequence with the catheter, enabled by having cooling in place.[43, 44]

To date, no AEF has yet been identified in a patient treated with active esophageal cooling using a dedicated esophageal cooling device, and only a single pericardio-esophageal fistula has been reported despite over 22,000 RF catheter ablations now completed using this cooling device.[45] Pericardio-esophageal fistula is a rare, and less severe, subset of fistula formation.[46] Analysis of this event suggests higher total energy was deposited over the esophagus than is typical, with higher ablation index targets on the posterior wall than the commonly used 380-400 units, and lesions stacked near the esophagus, which has recently been suggested to increase risk of injury.[33, 47-51] This may serve to emphasize that, like most technologies employed for safety, there may be limitations in the protective capabilities of cooling. Complications from the esophageal cooling device itself have been infrequent, with most occurring during long-term use for critical care patients (where durations of use extend well beyond 24 hours).[52-54]

New technologies may arrive that further reduce the risk of AEF, such as irreversible electroporation, or pulsed field ablation (PFA); however concerns remain about both the long-term efficacy of procedures utilizing PFA,[55, 56] as well as growing awareness of risks emerging with PFA, including stroke, acute coronary artery spasm, and delayed arterial injury via neointimal hyperplasia and medial fibrosis resulting in arterial stenosis.[57-59] Moreover, reports of fistula formation after irreversible electroporation are abundant in the oncology literature, which raises the question of whether PFA can actually eliminate the risk of AEF.[60-68] As such, the importance of technologies that may reduce this dreaded complication remains high.

## Limitations

The specific approach to data collection at each site was not standardized, and different methods of determining event rates at different sites were inevitable; however, at each site, the same approach was used for the entirety of the time frame studied (both before and after adoption of cooling), which may serve to limit significant bias in the data. In three of the 25 hospital systems included in this analysis, some physician operators did not adopt esophageal cooling at the same time, did not use cooling with the same consistency, or had not yet adopted cooling. All cases for which active esophageal cooling was used, as well as all cases of AEF regardless of adoption status, were nevertheless included in this analysis. Additional factors that could influence the rate of AEFs have not been explicitly quantified in this analysis; however, the time frames included are recent enough that major changes in technology or ablation technique have not occurred. Operators have generally been using predominantly higher power, shorter duration ablation settings, and using contact-force sensing irrigated radiofrequency ablation catheters with the same Ablation Index or Lesion Index targets during the time frame evaluated. Likewise, over this time frame, no significant secular trend in AEF rates has occurred.[10-13, 69, 70] Moreover, comorbidities such as congestive heart failure, coronary artery disease, lung disease, chronic kidney disease, and age have increased significantly among patients undergoing AF ablation,[5] which may further increase the likelihood of adverse events in the more recently treated population (which includes those in the post-adoption cohort that received esophageal cooling).[7, 71] Operator-specific practice changes, particularly after experiencing an AEF, may alter the risk of a subsequent AEF; however, most sites (64%) in this study had not experienced an AEF before adopting cooling.

## Conclusion

Adoption of active esophageal cooling during RF ablation of the left atrium for the treatment of atrial fibrillation was associated with a significant reduction in AEF rate.

## Data Availability

Data are available upon reasonable request.

